# Characterisation of placental, fetal brain and maternal cardiac structure and function in pre-eclampsia using MRI

**DOI:** 10.1101/2023.04.24.23289069

**Authors:** Megan Hall, Antonio de Marvao, Ronny Schweitzer, Daniel Cromb, Kathleen Colford, Priya Jandu, Declan P O’Regan, Alison Ho, Anthony Price, Lucy C. Chappell, Mary A. Rutherford, Lisa Story, Pablo Lamata, Jana Hutter

## Abstract

**Background:** Pre-eclampsia is a multiorgan disease of pregnancy that has short- and long-term implications for the woman and fetus, whose immediate impact is poorly understood. We present a novel multi-system approach to MRI investigation of pre-eclampsia, with acquisition of maternal cardiac, placental, and fetal brain anatomical and functional imaging.

**Methods:** A prospective study was carried out recruiting pregnant women with pre-eclampsia, chronic hypertension, or no medical complications, and a non-pregnant female cohort. All women underwent a cardiac MRI, and pregnant women underwent a fetal-placental MRI. Cardiac analysis for structural, morphological and flow data was undertaken; placenta and fetal brain volumetric and T2* data were obtained. All results were corrected for gestational age.

**Results:** Seventy-eight MRIs were obtained during pregnancy. Pregnancies affected by pre-eclampsia demonstrated lower placental and fetal brain T2*. Within the pre-eclampsia group, three placental T2* results were within the normal range, these were the only cases with normal placental histopathology. Similarly, three fetal brain T2* results were within the normal range; these cases had no evidence of cerebral redistribution on fetal Dopplers. Cardiac MRI analysis demonstrated higher left ventricular mass in pre-eclampsia with 3D modelling revealing additional specific characteristics of eccentricity and outflow track remodelling.

**Conclusions:** We present the first holistic assessment of the immediate implications of pre-eclampsia on the placenta, maternal heart, and fetal brain. As well as having potential clinical implications for the risk-stratification and management of women with pre-eclampsia, this gives an insight into disease mechanism.

**What is new?:**

- We propose a comprehensive MRI protocol for maternal cardiac and fetal-placental imaging that is safe in pregnancy and acceptable to women, and report on the largest set of functional placental MRI data in pre-eclamptic pregnancies
- The findings suggest altered oxygenation and microstructure in the placenta, which is associated with similar changes in the fetal brain
- The study identifies a specific pattern of remodelling in the maternal left ventricle that is associated with pre-eclampsia

**What are the clinical implications?:**

- We demonstrate that MRI provides maternal and fetal insights into pre-eclamptic pregnancies that cannot be obtained by other means during pregnancy
- Functional placental MRI may be able to differentiate between preeclampsia disease severities, while fetal brain imaging may help refine neurodevelopmental prognostic assessment of children exposed to pre-eclampsia in utero
- A specific 3D pattern of left ventricular remodelling and thickening was found to be discriminant of the presence of pre-eclampsia and may provide a basis for improved clinical risk stratification

## Introduction

Pre-eclampsia occurs when placental malperfusion, and a resultant release of soluble factors into the circulation, causes maternal vascular endothelial injury with subsequent hypertension and multi-organ damage.^1^ The Incidence is between 2 and 8%, with geographical variation.^2^ Globally, 14% of maternal deaths are associated with hypertensive diseases of pregnancy,^3^ which can occur secondary to multiple complications including renal and hepatic failure, stroke, eclampsia, pulmonary oedema, and disseminated intravascular coagulopathy. Cardiac manifestations include reduced cardiac output,^4^ increased vascular resistance, left ventricular hypertrophy,^5^ and diastolic dysfunction.^6^ After the postnatal period, women who have had pre-eclampsia remain at an increased risk of vascular disease and ischaemic heart disease.^7, 8^ While it is well recognised that maternal biochemical abnormalities can be detected prior to clinical onset of pre-eclampsia, there is also some evidence that cardiovascular maladaptation precedes the diagnosis of pre-eclampsia: echocardiography has demonstrated abnormalities in left ventricular mass in the late third trimester prior to the development of term pre-eclampsia.^9, 10^

Cardiac magnetic resonance imaging (MRI) combines excellent spatial and temporal resolution, and is the gold-standard non–invasive method of assessing cardiac morphology and function. In contrast with echocardiography, cardiac MRI is not limited by geometric assumptions in assessing ventricular volumes and image acquisition is less operator- dependent. This allows highly reproducible and accurate assessment of cardiac structure and function,^11^ making it the ideal modality for assessing differences between small groups.^12^ We have provided pilot data demonstrating feasibility of cardiac MRI in pregnancy, but large prospective studies are still required.^13^

For the fetus, pre-eclampsia is associated with fetal growth restriction (FGR) and complications such as placental abruption; excluding congenital anomalies, 5% of stillbirths occur in women with pre-eclampsia (Cantwell et al, 2011). In infants born to women with pre-eclampsia, increased rates of hypertension and cardiac dysfunction, stroke, cognitive dysfunction and psychiatric morbidity have been demonstrated.^14^

Postnatally obtained placental histopathological correlates with pre-eclampsia include both villous and vascular lesions,^15^ but sufficient *in vivo* and *in vitro* modelling of placental development in disease is lacking. MRI offers an opportunity for *in vivo* study of the placenta in pre-eclampsia, as well as evaluation of maternal and fetal structures. Recent studies applying functional MRI techniques to the placenta have proposed reduced oxygenation in the pre-eclamptic placenta via demonstration of increased heterogeneity and decreased mean T2*.^16, 17^ There is evidence of decreased placental diffusivity in small for gestational age fetuses.^18, 19^ Furthermore, advances in acquisition and reconstruction techniques have allowed more detailed investigation of the fetal brain.

Here we describe a multi-system approach to MRI investigation of pre-eclampsia, with acquisition of maternal cardiac, placental, and fetal brain anatomical and functional imaging.

## Methods

A prospective observational study was performed between 2019 and 2022 at Guy’s and St Thomas’ NHS Foundation Trust (London Dulwich Ethics Committee 08/LO/1958). Additional high-risk placental and fetal datasets were obtained from a previous study (London Dulwich Ethics Committee 16/LO/1573). Additional non-pregnant control datasets were obtained from a previous study (London - West London & GTAC Research Ethics Committee 17/LO/0034).

Women were recruited into four categories: non-pregnant controls, pregnant controls, women with pre-eclampsia, and pregnant women with chronic hypertension; a fifth category ‘complicated pregnancies’ was created retrospectively for women who had been recruited as controls but had developed other pregnancy complications and this group was excluded from further analysis. Women with diabetes and a hypertensive disorder were analysed within the appropriate hypertensive cohort. For all groups inclusion criteria included a body mass index (BMI) <40kg/m^2^ (with the MRI bore size the limiting factor), aged 16-50 years, and no contraindication to MRI such as non-compatible metal implants or claustrophobia. All women could undergo up to three scans.

Inclusion criteria for the control group was women with no previous cardiovascular or inflammatory cardiac history. Pregnant controls were excluded if they had a multi-fetal pregnancy or fetal congenital anomalies. Pregnant controls were moved into the complicated pregnancy cohort if they developed any other complications of pregnancy, had an infant born preterm, below the 3^rd^ centile, above the 97^th^ centile, or that required neonatal intensive care for any reason. Additionally, age and ethnicity matched non-pregnant controls, having undergone equivalent imaging, were recruited from the UK Digital Heart Project (UKDHP) at Imperial College London. Enrolment into this study excluded participants who had known cardiovascular or metabolic disease, as previously described.^20^

Case inclusion criteria was a diagnosis of pre-eclampsia (including superimposed on chronic hypertension) defined by the International Society for the Study of Hypertension in Pregnancy as hypertension with one of the following new-onset conditions at 20 weeks’ gestation or beyond: proteinuria, acute kidney injury, hepatic dysfunction, neurological features, haemolysis or thrombocytopaenia, or fetal growth restriction (Brown et al, 2018). Women with chronic hypertension were recruited as a separate cohort.

Demographic and clinical information, including on previous obstetric history, was collected on all participants. Neonatal outcomes including gestation at birth, mode of delivery, condition at birth, need for neonatal support, and birthweight were collected.

### MRI

After informed consent was obtained, participants had an MRI on a clinical 1.5T Philips Ingenia scanner using a combined 24-channel posterior and torso (dStream) coil. Maternal comfort in the supine position was achieved by careful positioning of the head and legs on elevated cushions, back padding, and additional cushions as requested. The MRI was performed in two sessions (maternal cardiac, and fetal-placental) each of which lasted around 30 minutes. A break was given in the middle of the session. In order to reduce any bias that could arise from maternal anxiety at the start of the scan, women with an odd case study ID underwent fetal-placental imaging first, and those with an even ID underwent cardiac imaging first.

An obstetrician or midwife was present for all scans. Maternal temperature was recorded before and after the scan. Continuous oxygen saturations were measured, and blood pressure was taken every 10 minutes.

The protocol is illustrated in Figure 1.

**Figure 1:**
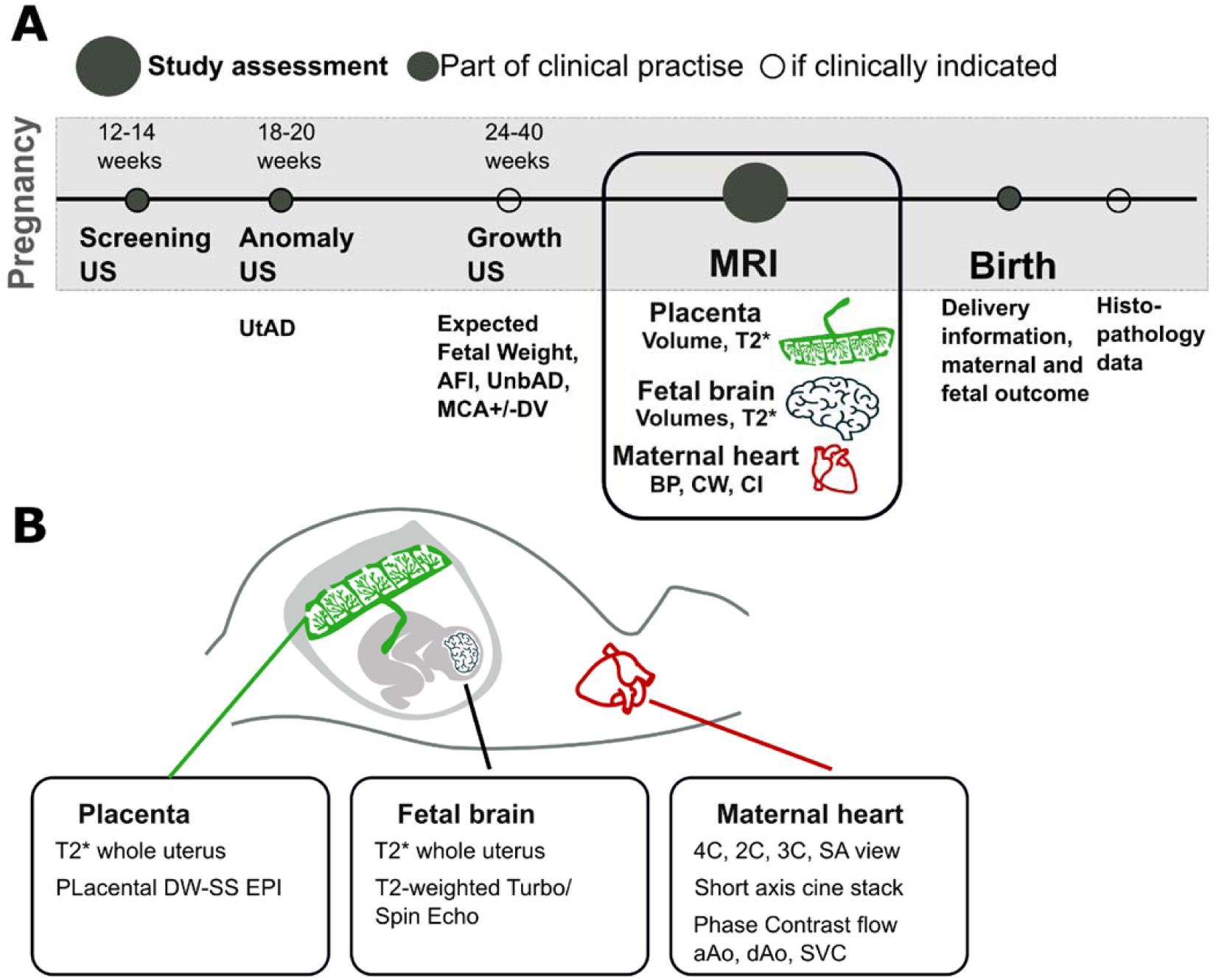
Schematic of study protocol. US: ultrasound; UtAD: uterine artery Doppler; AFI: amniotic fluid volume; UmbAD: umbilical artery Doppler; MCA: middle cerebral artery Doppler; DV: ductus venosus Doppler; BP blood pressure; CW cardiac work; CI: cardiac index; DW-SS EPI: diffusion weighted single shot echo planar imaging; 4C: 4 chamber; 2C 2 chamber; 3C 3 chamber; SA: short axis; aAo ascending aorta; dAo descending aorta; SVC superior vena cava.

### Fetal-placental MRI

Fetal brain and whole uterus structural imaging was performed in five planes (transverse centred on fetal brain; two opposing sagittal oblique planes; maternal coronal plane and maternal sagittal plane) in order to obtain whole uterus and dedicated placental images. MRI was undertaken using 2D-single-shot Turbo Spin Echo (resolution 1.5x1.5x2.5mm, field of view (FOV) 320x320x110mm, echo time (TE) 180ms). Fetal body imaging was performed for clinical reporting only.

Placental T2* mapping was obtained using a multi-echo gradient-echo sequence (2.5mm isotropic, FOV=300x300x110mm, TE=11, 58, 117, 176ms). Placental diffusion-weighted imaging used a single-shot echo-planar-imaging sequence with parameters adapted to meet the expected diffusivity (1 b=0, 6 b=375, 6 b=750; transverse relaxation (TR)=6.6ms, TE=78ms, matrix = 512x512x56, resolution 2x2x4mm). Both sequences were repeated twice to improve data robustness.

### Cardiac MRI

Anatomical and functional cardiac MRI sequences were acquired using commonly employed sequences adapted to reduce acoustic noise and heating, and so improve suitability in pregnancy. Views taken included: 4 chamber, 2 chamber, 3 chamber, short axis and outflow (acquired using dynamic balanced stead state free precession sequences with 30 cardiac phases, 11 second breath-hold, FA=60, TE/TR∼2/4ms, resolution=1.7x1.7x8mm); and a short axis stack (14 slices, sense=2, resolution 2x2x10mm). Phase contrast flow sequences of the ascending and descending aorta, and the superior vena cava were acquired with individually adapted encoding velocities (30 phases, 14 second breath-holds, sense=2, resolution=2.5x2.5x8mm). A pragmatic decision was made to obtain inferior vena cava images during the ‘fetal-placental’ portion of the MRI as the coil was appropriately placed. The scanning parameters are summarised in Table 1. Images acquired are demonstrated in Figure 2.

**Figure 2:**
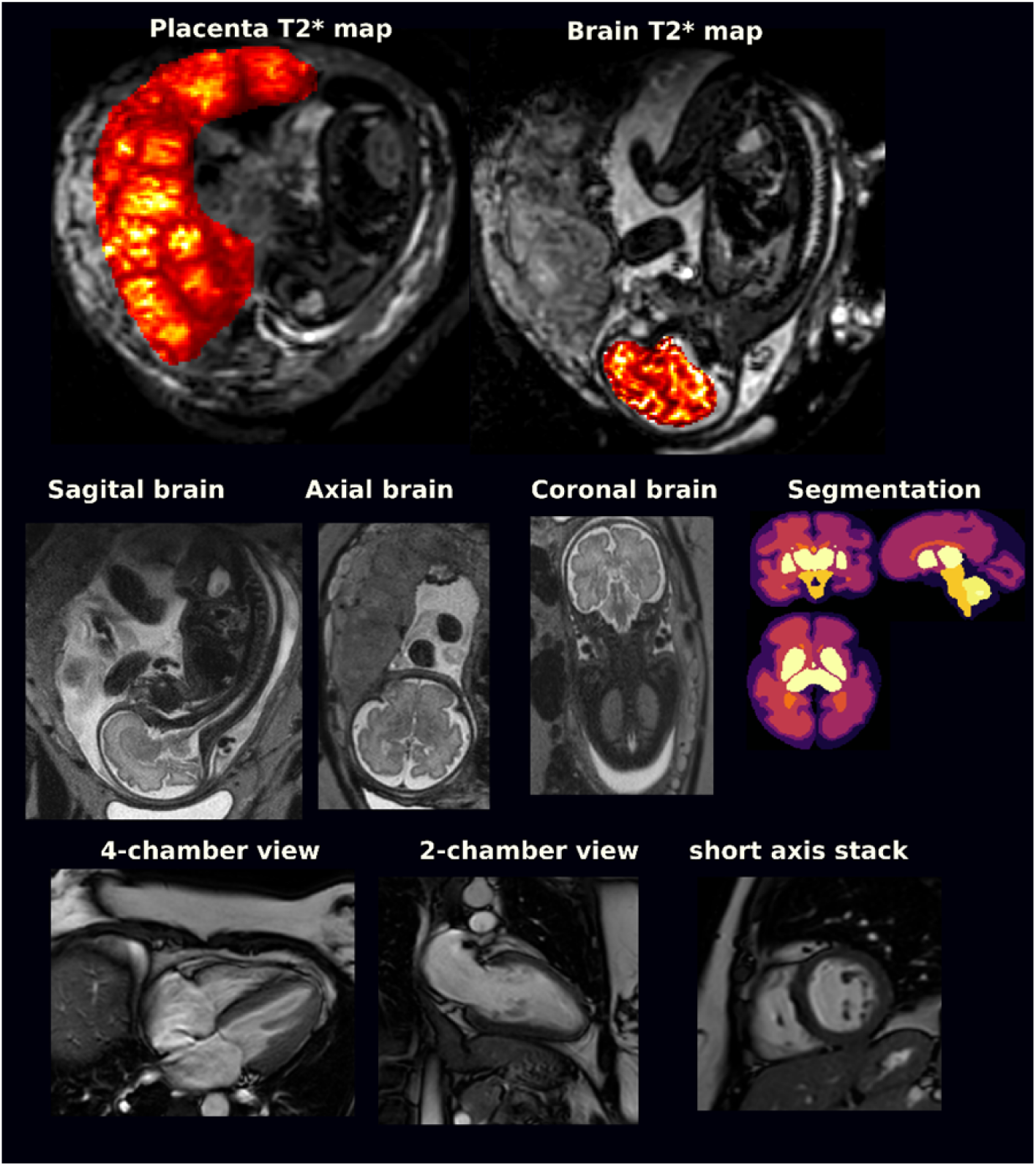
Summary of images acquired on protocol.

**Table 1:** Scanning parameters

| **Placental and Fetal MRI** |  |
| --- | --- |
| **Anatomical** | **T2 weighted Turbo Spin Echo**<br>TR 30s; TE 180ms; Matrix size 384x384x150;<br>Resolution 1.25x1.25x2.5mm |
| **T2\* static** | **Multi-Echo Gradient Echo**<br>TR 23s; TE<br>12.599ms/65.34ms/118.08ms/170.821ms/223.562ms;<br>Dynamics – 2; Matrix size 224x224x850; Resolution<br>2.5x2.5x2.5mm |
| **Diffusion** | **Pulsed Gradient Echo Spin Echo, Echo Planar Imaging**<br>TR 6.6s; TE 78ms; Matrix 512x512x56; Resolution<br>2x2x4mm; b 375 (6 directions); b 750 (6 directions) |
| **T2\* dynamic** | **Multi-Echo Gradient Echo**<br>TR 6.4s; TE 7.84ms/60.574ms/113.308ms/116.041ms;<br>Dynamics – 30; Matrix size 256x256x360; Resolution<br>2.5x2.5x2.5mm |
| **Cardiac MRI** |  |
| **Cine bSSFP** | **bSSFP**<br>30 phases; FA 60; TR 4ms; TE 2ms; Resolution<br>1.7.1.7.8mm; ~11 second breath-hold |
| **Short axis stack** | **bSSFP**<br>TR 4.1s; TE 2ms; 14 slices; Sense 2; Resolution<br>2x2x10mm |
| **Phase contrast (breath-hold option)** | 30 phases; Sense 2; Resolution 2.5x2.5x8mm; 14<br>second breath-hold |
| **Phase contrast (free breathing option)** | 30 phases; Sense 2; Resolution 2.5x2.5x5mm; NSA 4 |

*TR: repetition time; TE echo time; bSSFP balanced Steady State Free Precession*

Non-pregnant controls from the healthy control group underwent an equivalent cardiac MRI protocol on a 1.5-T Philips Achieva system (Best, the Netherlands) as previously described.^20^

### Analysis

The placenta was visually inspected on a T2 image. Manual outlining was performed on a functional scan mapped to subsequent scans. Monoexponential fitting was performed to obtain T2* maps. Mean placenta T2*, histogram skewness and kurtosis, and mean diffusivity (as the apparent diffusion coefficient – ADC) were calculated.

Brain volume (supratentorial tissues excluding brainstem and cerebrospinal fluid) was manually segmented on functional maps. Mean T2* values were obtained by averaging the T2* maps from manually outlined brains.

The cardiac MRIs were analysed using cvi42 post-processing software (Version 5.1.4, Circle Cardiovascular Imaging Inc., Calgary, Canada), using standard clinical methodology, by operators masked to the clinical diagnosis.^21^

The 3D left ventricular morphology was studied by the construction of a statistical shape model from the segmentations of the short axis stack at the end diastolic frame as reported in previous studies.^22–24^ Briefly, 3D meshes were built using a computational anatomy tool kit,^25^ anatomical modes of variation were found by Principal Component Analysis, and the impact of pre-eclampsia was assessed by an optimised linear discriminant analysis of the first 12 modes.

Flow data was processed using CVI42 Flow version 5.10.3. The magnitude image with the sharpest contrast was used to determine vessel contours. Contours were then propagated to phase contrast images (with manual correction as required) in all temporal phases. All parameters were first assessed against gestational age, and then as cases against controls. Matching was undertaken to nearest gestational age, with further analysis using the Mann-Whitney U Test.

### Statistical Analysis

Statistical analysis was performed with R version 3.6.0 (R Foundation for Statistical Computing) and RStudio Server version 1.043, unless otherwise stated. Variables are expressed as percentages if categorical, mean ± SD if continuous and normal, and median (interquartile range) if continuous and non-normal. Baseline anthropometric data were compared by using Kruskal-Wallis tests and, if differences were identified, a Wilcoxon test was used for pairwise comparisons with Benjamini-Hochberg adjustment for multiple testing. Imaging parameters in 2 or more groups were compared by using analysis of covariance, adjusted for relevant clinical covariates. When differences were significant, a Tukey post hoc test was applied for pairwise multiple comparisons.

## Results

### Study participants

Seventy-eight scans were obtained from 65 pregnant women (with nine women having two scans, and two having three scans). An additional 38 scans were obtained from healthy non-pregnant controls. Cardiac assessment was not completed in two pregnant participants: one owing to claustrophobia, and one to worsening hypertension. For a further 16 women, no cardiac data were obtained. Nine women developed pregnancy complications after being scanned as a control and so were moved to the complicated pregnancy cohort and excluded. Thirteen women with pre-eclampsia were recruited, one of whom had three MRIs. Of the 13, six had pre-eclampsia superimposed on chronic hypertension. Table 2 summarises the demographics of all cohorts. Supplementary Table 1 details relevant obstetric history and outcomes of the pre-eclamptic cohort.

**Table 2:** Demographics of participants

| **Cohort (number of scans)** | **Pregnant controls (n=32)** | **Non-pregnant controls (n=38)** | **Pre-eclampsia (n=15)** | **Chronic hypertension (n=22)** | **Complicated pregnancy (n=9)** |
| --- | --- | --- | --- | --- | --- |
| **Age (years), median (IQR)** | 34.4<br>(31.8-37.3) | 32.0<br>(27.0-39.0) | 33.5<br>(30.8-37.8) | 36.8<br>(33.8-41.5) | 33.1<br>(31.9-35.3) |
| **BMI (kg/m<sup>2</sup>), median (IQR)** | 24.7<br>(22.9-27.2) | 25.3<br>(21.2-25.4) | 28.1<br>(24.1-33.2) | 31.9<br>(27.4-33.8) | 23.1<br>(21.9-24.4) |
| **Gestational age at the time of scan (weeks), median (IQR)** | 28.1<br>(23-5-32.1) |  | 32.7<br>(30.0-34.1) | 27.8<br>(25.0-32.0) | 27.3<br>(24.4-33.3) |
| **Gestational age at birth, mean (SD)** | 39.6<br>(1.07) |  | 33.4<br>(2.77) | 37.8<br>(0.7) | 36.3<br>(2.87) |
| **Birth weight (grams), mean (SD)** | 3394<br>(402) |  | 1781<br>(552) | 2973<br>(636) | 2370<br>(978) |
| **Birth weight (grams), range** | 2710-4175 |  | 695-3070 | 1560-4840 | 1010-3220 |
| **Previous pre-eclampsia (%)** | 0 | 0 | 3 (25) | 2 (8) | 0 |

*BMI: body mass index; IQR: interquartile range*

### Placenta

In the cross-sectional control group, there was a decrease in mean T2* values across gestation (p<0.01). This was also seen as a trend (not reaching significance) in the chronic hypertensive group. The pre-eclamptic cohort had significantly lower T2* values throughout gestation, and a gradual decline was not seen. Both skewness and kurtosis increased throughout gestation in the control group, with the chronic hypertensive group overlying this, although not reaching significance. The pre-eclamptic group had higher skewness and kurtosis than both other groups, and placental volume was reduced (p=0.02) (Figure 3). Three women with pre-eclampsia had placental T2* values similar to controls; of note these were the only three women without any vascular abnormalities on placental histopathology (Table 3, Supplementary Table S1).

**Figure 3:**
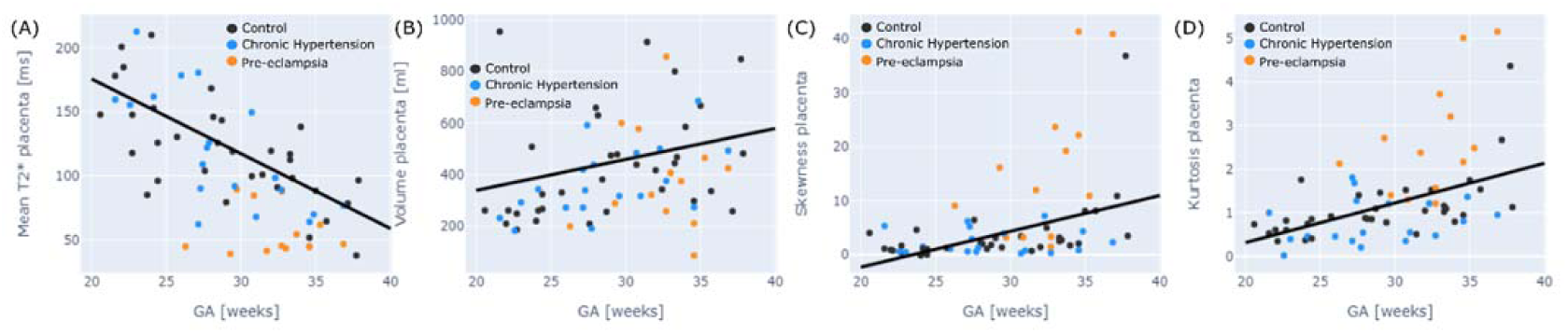
Placental anatomical and T2* by group. Figure 1: A) mean placental T2* by gestation; B) placental mass by gestation; C) skewness and D) kurtosis by gestation. Black dot = healthy pregnancy; blue = chronic hypertension; orange = pre-eclampsia.

**Table 3:** Relationship between fetal brain and placental T2* and clinical parameters

| Participant ID | Placental T2* | Placental histopathology | Fetal brain T2* | Fetal Dopplers | Fetal growth restriction |
| --- | --- | --- | --- | --- | --- |
| 1 | Low | Abnormal | Low | Redistribution | Yes |
| 2 | Low | Other vascular abnormality | Low | Normal | Yes |
| 3 | Low | Abnormal | Low | Redistribution | Yes |
| 4 | Low | Abnormal | Low | Redistribution | No |
| 5 | Normal | Normal | Normal | Normal | No |
| 6 | Low | Abnormal | Low | Redistribution | Yes |
| 7 | Low | Abnormal | Low | Redistribution | Yes |
| 8 | Normal | No pre-eclampsia related changes | Normal | Normal | Yes |
| 9 | Low | Not available | Low | Redistribution | No |
| 10 | Low | Abnormal | Low | Redistribution | Yes |
| 11 | Low | Abnormal | Low | Redistribution | Yes |
| 12 | Low | Abnormal | Low | Redistribution | Yes |
| 13 | Normal | Normal | Normal | Normal | No |

Visual inspection of control versus pre-eclamptic placentas revealed a more variable lobule size and less consistent signal intensity in women with pre-eclampsia. There was also an increase in low signal areas as compared to controls.

### Fetal brain

Fetal brain T2* values were demonstrated to decline with advancing gestation among controls. This relationship was preserved in the chronic hypertensive group. Among women with pre-eclampsia, the fetal brain T2* values were significantly reduced through all gestations (p<0.01) and did not have a linear decline with gestation (Figure 4). There were three cases in which brain T2* was in line with controls; in all these cases fetal Doppler studies were normal (Table 3, Supplementary Table S1). One case demonstrated an abnormal T2* values but with preserved fetal Doppler studies (Table 3, Supplementary Table S1). Brain volume was reduced in the presence of pre-eclampsia (p=0.03) (Figure 4). Z scores were generated for mean placental T2* values and mean fetal brain T2* values in order to allow for direct comparison: there was a direct correlation between normal and low scores in both domains (Figure 5).

**Figure 4:**
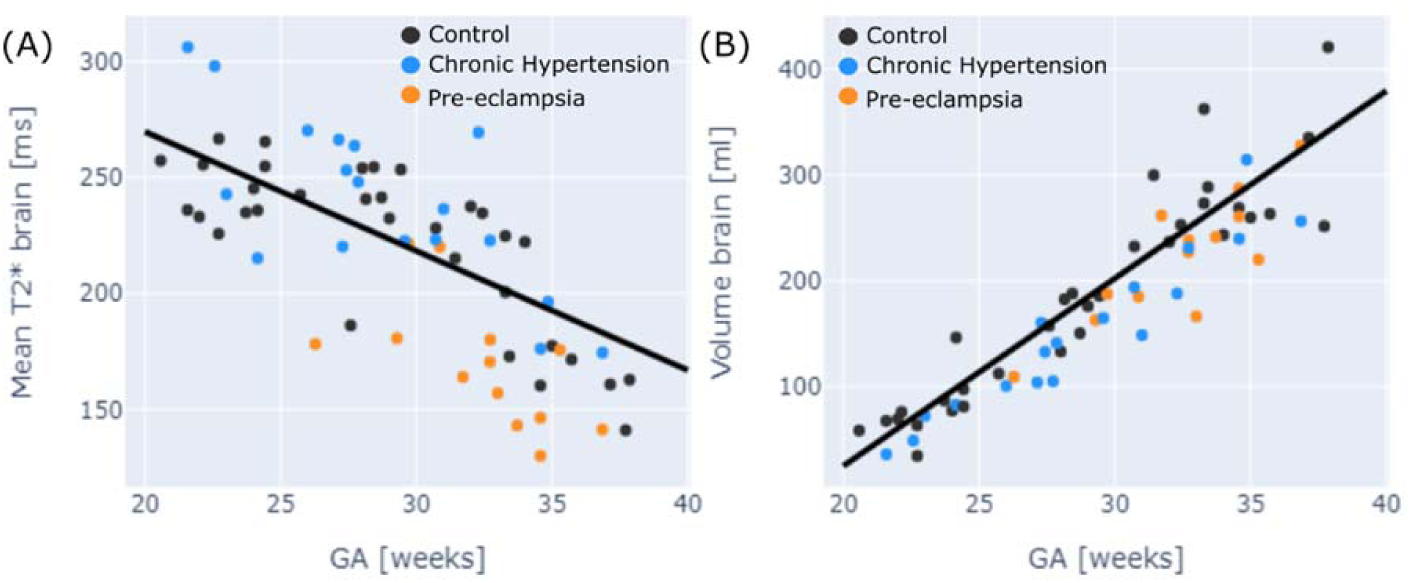
Fetal brain anatomical and T2* by group. Figure 2: A) Fetal brain mean T2* by gestation; B) fetal brain volume by gestation. Black dot = healthy pregnancy; blue = chronic hypertension; orange = pre-eclampsia.

**Figure 5:**
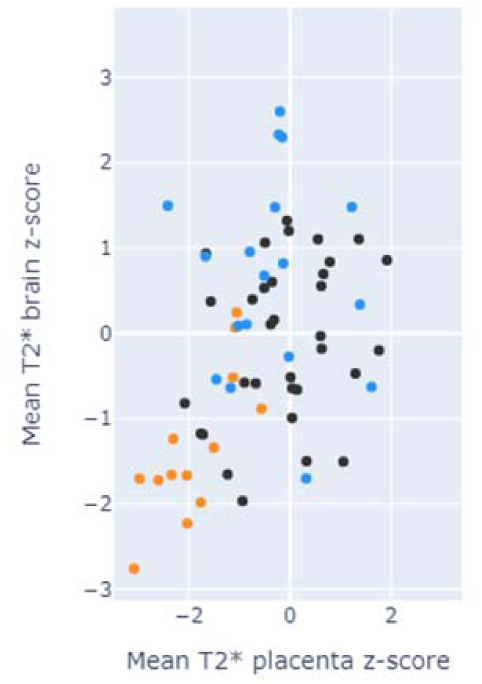
Comparison of fetal brain mean T2* z-score and placental mean T2* z-score. Black dot = healthy pregnancy; blue = chronic hypertension; orange = pre-eclampsia.

Major differences in the placental and fetal findings of the control and pre-eclampsia groups are summarised in Supplementary Figure 1.

### Cardiac MR

Systolic (p<0.001) and diastolic (p=0.03) blood pressure were lower in the pregnant controls (107 / 66 mmHg) than in pre-eclamptic participants (121 / 77 mmHg). There were no other baseline anthropometric differences between groups.

After adjusting for body surface area (BSA), non-pregnant controls had lower heart rate (68 bpm vs 74.8 bpm; p = 0.02), cardiac output (5.5 L / min vs 6.2 L/ min; p = 0.02), LV mass (77.4 g vs 87 g; p = 0.01) and concentricity (LV mass / LV end-diastolic volume; 0.57 g / ml vs 0.64 g / L; p = 0.01) than pregnant participants. The LV mass of the pre-eclamptic group (98.05 ± 24.5 g) was higher than both the pregnant controls (83.3 ± 12.08 g; p = 0.04) and the non-pregnant group (77.4 ± 13.8 g; p=0.04). The study of the 3D anatomy revealed a specific thickening pattern (existence of regional increase in wall thickness wall locations of mid antero-lateral and postero-septal) together with changes in eccentricity (the ventricular cross-section displayed a dilation in the axis oriented in the direction of the outflow tract) and the onset of a bulge below the outflow track, see Figure 3. A detailed inspection of the modes of anatomical variation further reveals that the thickening pattern associated to pre- eclampsia was linked to a localised basal concentric remodelling and not to the complementary basal eccentric remodelling (see modes 11 and 13 in Supplementary Figure S2).

## Discussion

### Summary of main findings

We have demonstrated a comprehensive anatomical and functional MRI protocol for cardiac and fetal-placental imaging that is safe in pregnancy and acceptable to women, with 97% of scans completed. As far as we are aware, this is the largest set of functional placental MRI data in pre-eclamptic pregnancies, and confirms previous findings suggestive of altered oxygenation and microstructure. We have demonstrated altered T2* in the fetal brain, in proportion to that seen in the placenta. Finally, we have characterised the remodelling pattern of the maternal left ventricle associated with the presence of pre-eclampsia. Conventional cardiac MRI analysis demonstrated higher LVM in pre-eclampsia than in uncomplicated pregnancies, while 3D statistical modelling revealed specific characteristics of eccentricity and outflow track remodelling beyond the thickening of the walls.

### Strengths and limitations

This is the largest study to date investigating maternal, fetal and placental implications of pre-eclampsia *in vivo*. We utilise an optimised protocol^26^ and includes novel findings related to the T2* values. To our knowledge this is the first published study making a holistic assessment of the fetal, placental and maternal implications of pre-eclampsia. The MRI sequence described here is safe in pregnancy and acceptable to women, and gives insight into pre-eclamptic pregnancies that cannot be obtained by other means during pregnancy. The use of consistent maternal positioning and scanning protocols means that the data are robust and comparable between subjects.

Functional placental data build on that previously presented to demonstrate abnormalities in microstructure and oxygenation in the pre-eclamptic placenta.^17, 18^ The inclusion of a non-pregnant cardiac control group delineates changes expected in pregnancy from those that are pathological. Motion correction and reconstruction of fetal images has allowed for the first analysis of T2* brain maps in fetuses affected by pre-eclampsia. The inclusion of a chronic hypertensive group demonstrates where phenotypes are driven by pre-eclampsia, rather than the gestational effects of hypertension alone.

There are several limitations to this study: firstly, recruitment to the pre-eclampsia group was challenging, largely due to the often short intervals from diagnosis to delivery and so the study is relatively small, perhaps obscuring other significant differences between cases and controls. Secondly, the lack of long-term follow up limits understanding of how much the observed maternal cardiac and fetal brain changes directly impact chronic health outcomes. Thirdly, the small sample size makes it difficult to comment on confounders within the pre-eclampsia group, in particular the presence of diabetes, and early versus late onset disease.

### Comparison to other work

The placental phenotype in control pregnancies is in line with previous work: the reduction in T2* across gestation in normal pregnancies has been demonstrated using both our imaging protocols^26^ and those that of others.^27, 28^ As previously demonstrated, there is overlap between chronic hypertensive pregnancies and normal pregnancies in terms of mean T2*, skewedness and kurtosis.^16^ Pregnancies affected by pre-eclampsia are typically outliers in all domains. This is to be expected given the higher rate of hypoxic vascular anomalies seen in the pre-eclamptic placentae compared to that affected by chronic hypertension alone.^29^

In all cases where the placental T2* in pre-eclamptic pregnancies lay within the normal range, there was normal placental histopathology and delivery at later gestations, suggesting a less severe clinical phenotype. Pre-existing work on placental perfusion MRI in women with early and late pre-eclampsia, where women with later onset disease has demonstrated preserved placental perfusion.^30^ The decreased placental perfusion seen in the early pre-eclampsia group, as well as histopathological evidence for greater maternal malperfusion in early onset pre-eclampsia as compared to late,^31^ provide basis for the variation seen in our cohort. Our findings go further to support the ability of functional placental MRI to differentiate between disease severities.

Novel to this study is the association between placental T2* and fetal brain T2*. The negative correlation between decreasing brain T2* and increasing gestation in the normal pregnancy is already established, and thought to be secondary to a number of structural changes including decreasing water density and increasing myelination.^32^ Preservation of brain volume with a decrease in T2* in the pre-eclampsia group may reflect a reduction in oxygenation, with or without reduction in angiogenesis. This seems particularly likely given that this does not seem to occur in the absence of reduced placental T2*, and that in all but one case there is corresponding Doppler evidence of cerebral redistribution. Long-term neurodevelopmental and behavioural outcomes of children born to mothers with pre- eclampsia are recognised, and a small study has shown differences in neuroanatomy persisting at 7-10yrs;^33^ although this is difficult to disentangle from the effects of prematurity, our findings may suggest an antenatal antecedent.

Looking into the mother’s health, novel to this study is the 3D thickening pattern of the left ventricle that is found discriminant of the presence of pre-eclampsia. The extra workload in pre-eclampsia is indeed increasing the cardiac mass and mass to volume ratio, as previously reported in much larger echocardiography studies.^5^ This demonstrates the higher power of cardiac MRI to identify differences between small groups, and its role in cardiovascular research in pregnancy. Our preliminary data further reveal an intriguing pattern of thickening, eccentricity and bulging as illustrated in Figure 6. The fact that the other groups of non-pregnant controls and chronic hypertension did not show differences with control pregnancy along this axis suggests that this is an acute maladaptive remodelling process specific to pre-eclampsia. In relationship with this finding, the chronic remodelling of the left ventricle 5 to 10 years after a hypertensive pregnancy manifest as a global concentric (and not eccentric) thickening of the left ventricle.^34^ Intriguingly in the response of pre-eclampsia during pregnancy it is only the base of the heart where this concentric (and not eccentric) thickening pattern is seen (see Supplementary Figure S1). This collection of observations leads us to hypothesize that it is the base of the heart the region that is first recruited and most intensively working in response to pre-eclampsia, and that the rapid onset of hypertension causes a level of uneven thickening pattern and septal bulging.

**Figure 6:**
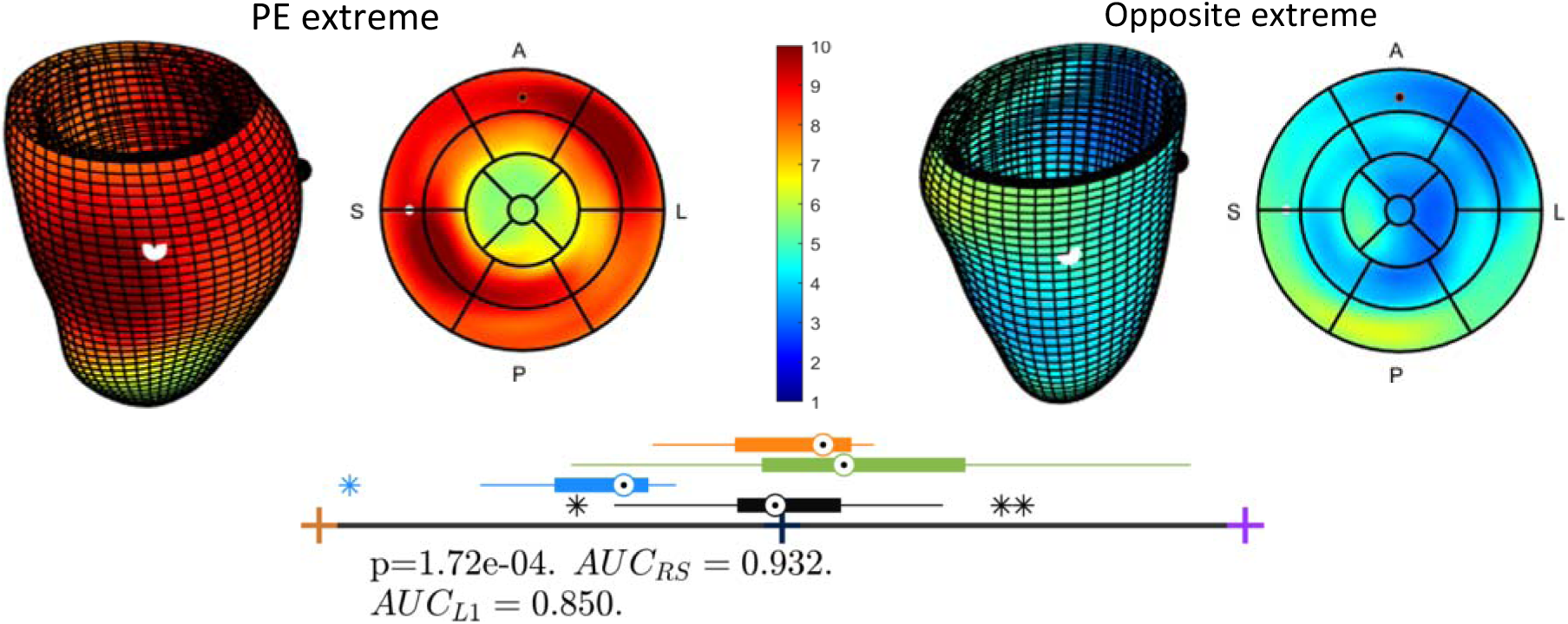
Left ventricular remodelling associated with pre-eclampsia. Figure 3: Left ventricular remodelling associated with pre-eclampsia (PE), illustrating thickened walls and eccentricity & bulging located at the outflow track. **Top panel:** extremes of the anatomical mode that best discriminate pregnant controls to pregnant pre-eclamptic subjects - The 3D and bullseye views are colour-coded by thickness (in mm) and have black and white spheres at the sides of the outflow track for easy mapping between both views – A: Anterior; P: Posterior; L: Lateral; S: Septal. **Bottom panel:** box-plot of distributions of pregnant controls (black), pre-eclampsia (blue), chronic hypertension (orange) and non- pregnant controls (green), together with metrics of the discriminant performance of the linear discriminant analysis between pregnant controls and pre-eclampsia (AUC: Area Under the Curve; RS: Resubstitution; L1: Leave-1 out cross-validation).

### Implications

While clinical fetal MRI is largely confined to assessment of structural anomalies, there is evidence here of a potential role for other high risk obstetric conditions. We have demonstrated feasibility of a comprehensive fetal, placental and maternal cardiac MRI in terms of safety, data acquisition and acceptability to women.

Future work into determining risk of developing, likely clinical severity and implications of pre-eclampsia should combine MRI, obstetric ultrasound and biomarkers, as it is likely that a combination of investigations could yield greatest information.

While we have demonstrated a correlation of findings associated with pre-eclampsia, further work should be done in defining this phenotype. As well as increasing the cohort size, attention should be paid to early and late onset disease, as these are likely to be mechanistically different and have different clinical end point.^35^ While fetal growth restriction has previously been associated with reduced placental T2*,^19,36^e we have been unable to delineate the implications of pre-eclampsia on this finding. Creation of a cohort where the interplay between these three findings could be investigated would be of value. MRI could have implications for earlier diagnosis of pre-eclampsia; high-risk women (for example those with early onset growth restriction) who are not yet diagnosed with pre- eclampsia should be included in future work, as this could have significant implications for their management. Moreover, the 3D specific remodelling pattern linked to pre-eclampsia could have implications for prognosis and risk models for future cardiovascular events. Any future work on cardiac imaging should include longitudinal and postnatal data in order to better delineate the mechanisms of continued cardiac risk, and for stratification of risk in individuals.

In terms of perinatal outcomes, we have demonstrated a relationship between pre- eclampsia and reduced fetal brain T2*, and also between reduced fetal brain T2* and fetal cerebral Doppler redistribution. While there is some evidence of abnormal neurocognitive outcomes in children who have been affected by redistribution (Beukers et al, 2017), further neurocognitive follow up of children with paired MRI and ultrasound data could improve understanding of the longer-term clinical implications of pre-eclampsia on the child.

In terms of techniques used, while the relationship between T2* and deoxyhaemoglobin is well established, it is subject to influence from other factors. Therefore, additional functional imaging, such as diffusion techniques, linked with recent advanced analysis techniques^37^ may lead to greater insight of the mechanisms of pre-eclampsia.

## Conclusions

MRI of pregnancies affected by pre-eclampsia reveals a reduction in mean placental and fetal brain T2*, with preservation of the volume of both organs. There is also an increase in maternal cardiac work and specific thickening patterns of the left ventricle. This all demonstrates that MRI is clinically valuable in risk stratification, investigation, and management planning of women at high risk of pre-eclampsia, or who have been diagnosed with the condition.

## Data Availability

The data that support the findings of this study are available from the corresponding author, MH, upon reasonable request.

## Non-standard abbreviations and acronyms

ADC: apparent diffusion coefficient
bpm: beats per minute
CO: cardiac output
EDV: end diastolic volume
ESV: end systolic volume
FGR: fetal growth restriction
FOV: field of view
GCS: global circumferential strain
GLS: global longitudinal strain
GRS: global radial strain
IQR: interquartile range
LV: left ventricular
LVM: left ventricular mass
TE: echo time
TR: repetition time

## Acknowledgments

We are grateful to the women who participated in this study.

---

**Supplementary Table S1:** Medical history and obstetric outcomes of the pre-eclamptic cohort.

|  |  |
| --- | --- |
| **Ethnicity (n)** |  |
| White | 2 |
| Black | 5 |
| South Asian | 4 |
| Other | 2 |
| **Parity (n)** |  |
| Nulliparous | 9 |
| Parous | 4 |
| **Gestational age at PET diagnosis (weeks) (range)** | 27.0 (20.7 – 35.0) |
| **Pre-eclampsia superimposed on chronic hypertension (n)** | 6 |
| **PIGF (pg/ml) (n)** |  |
| ≥12 | 2 |
| <12 | 2 |
| **Proteinuria (n)** | 13 |
| **Acute kidney injury (n)\*** | 5 |
| **Hepatic dysfunction (n)\*** | 8 |
| **Haemolysis (n)\*** | 1 |
| **Neurological involvement (n)\*** | 0 |
| **Thrombocytopaenia (n)\*** | 2 |
| **Fetal growth restriction (n)** | 10 |
| **Gestational age at MRI (weeks) (range)** | 32.0 (26.3 – 35.3) |
| **Antenatal aspirin use (n)** | 7 |
| **Antihypertensives at time of MRI (n)** |  |
| Single agent | 8 |
| Two agents | 4 |
| Three agents | 1 |
| **Maximum antihypertensives (n)** |  |
| Single agent | 5 |
| Two agents | 4 |
| Three agents | 3 |
| Four agents | 1 |
| **Evidence of redistribution on ultrasound at time of MRI (n)** |  |
| Yes | 9 |
| No | 4 |
| **Mode of delivery (n)** |  |
| Caesarean section in labour | 2 |
| Caesarean section prior to labour | 10 |
| Spontaneous vaginal | 1 |
| **Indication for delivery (n)** |  |
| Maternal | 4 |
| Fetal | 9 |
| **Neonatal complications (n)** |  |
| NICU admission | 12 |
| Respiratory support | 4 |
| Necrotising enterocolitis | 2 |
| Hypoglycaemia | 5 |
| Jaundice | 6 |
| **Placental histopathology (n)** |  |
| Abnormalities consistent with pre-eclampsia | 8 |
| Other abnormalities | 2 |
| Normal | 2 |
| Not available | 1 |

PIGF: placental growth factor.
\*Diagnosed at any stage in pregnancy or postnatal period

**Supplementary Figure S1:**
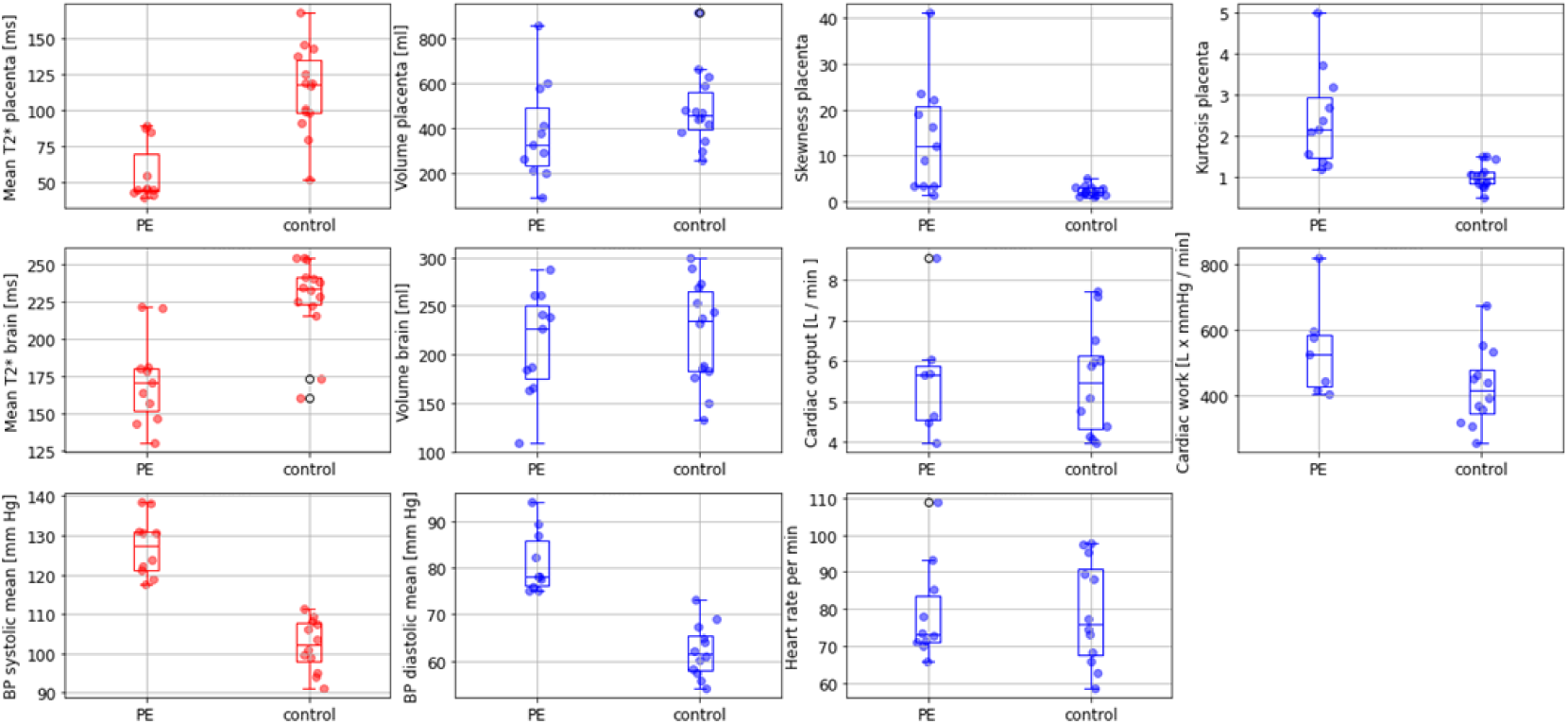
Summary of major findings in the control and pre-eclamptic cohorts. Placental mean T2* p<0.001; placental volume p=0.02; placental T2* skewness p=0.03; placental T2* kurtosis p=0.01. Fetal brain mean T2* p<0.001; fetal brain volume p=0.03. Cardiac output p=0.22; cardiac work p=0.905; mean systolic blood pressure p<0.001; mean diastolic blood pressure p=0.01; mean heart rate p=0.159 BP: blood pressure

**Supplementary Figure S2:**
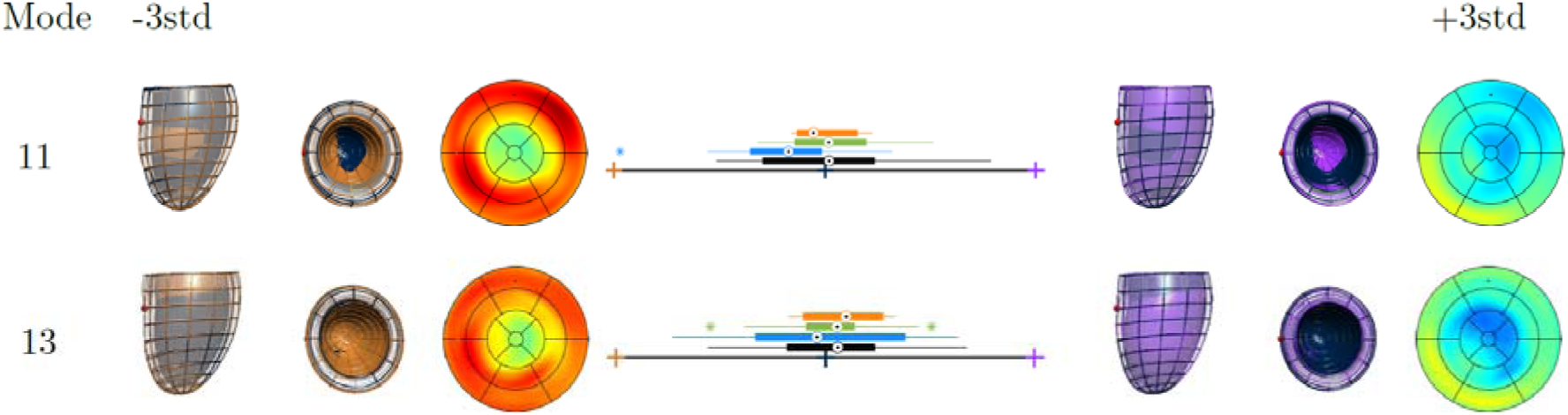
Anatomical models of variation that most correlate with thickness. Sup. Fig. 1: The two anatomical modes of variation that most correlated with thickness (R2=22.89% and 26.58% respectively for mode 11 and 13), with mode 11 showing a concentric basal thickening (narrower cavity at the base) and with mode 13 showing a complementary eccentric basal thickening (wider cavity at the base). Box-plots of the subjects (pregnant control: black; pre-eclampsia: blue; non-pregnant control: green; chronic hypertension: orange) show an almost significant difference in mode 11 between pregnant control and pre-eclampsia (p=0.07) but the complete overlap in mode 13 (p=0.77).

